# Multi-analyte proteomic analysis identifies blood-based neuroinflammation, cerebrovascular and synaptic biomarkers in preclinical Alzheimer’s disease

**DOI:** 10.1101/2024.06.15.24308975

**Authors:** Xuemei Zeng, Tara K. Lafferty, Anuradha Sehrawat, Yijun Chen, Pamela C. L. Ferreira, Bruna Bellaver, Guilherme Povala, M. Ilyas Kamboh, William E. Klunk, Ann D. Cohen, Oscar L. Lopez, Milos D. Ikonomovic, Tharick A. Pascoal, Mary Ganguli, Victor L. Villemagne, Beth E. Snitz, Thomas K. Karikari

**Author notes:** Corresponding author. Department of Psychiatry, School of Medicine, University of Pittsburgh, Pittsburgh 15213, PA, USA.

## Abstract

**Background:** Blood-based biomarkers are gaining grounds for Alzheimer’s disease (AD) detection. However, two key obstacles need to be addressed: the lack of methods for multi-analyte assessments and the need for markers of neuroinflammation, vascular, and synaptic dysfunction. Here, we evaluated a novel multi-analyte biomarker platform, NULISAseq CNS disease panel, a multiplex NUcleic acid-linked Immuno-Sandwich Assay (NULISA) targeting ∼120 analytes, including classical AD biomarkers and key proteins defining various disease hallmarks.

**Methods:** The NULISAseq panel was applied to 176 plasma samples from the MYHAT-NI cohort of cognitively normal participants from an economically underserved region in Western Pennsylvania. Classical AD biomarkers, including p-tau181, p-tau217, p-tau231, GFAP, NEFL, Aβ40, and Aβ42, were also measured using Single Molecule Array (Simoa). Amyloid pathology, tau pathology, and neurodegeneration were evaluated with [11C] PiB PET, [18F]AV-1451 PET, and MRI, respectively. Linear mixed models were used to examine cross-sectional and Wilcoxon rank sum tests for longitudinal associations between NULISA biomarkers and AD pathologies. Spearman correlations were used to compare NULISA and Simoa.

**Results:** NULISA concurrently measured 116 plasma biomarkers with good technical performance, and good correlation with Simoa measures. Cross-sectionally, p-tau217 was the top hit to identify Aβ pathology, with age, sex, and *APOE* genotype-adjusted AUC of 0.930 (95%CI: 0.878-0.983). Fourteen markers were significantly decreased in Aβ-PET+ participants, including TIMP3, which regulates brain Aβ production, the neurotrophic factor BDNF, the energy metabolism marker MDH1, and several cytokines. Longitudinally, FGF2, IL4, and IL9 exhibited Aβ PET-dependent yearly increases in Aβ-PET+ participants. Markers with tau PET-dependent longitudinal changes included the microglial activation marker CHIT1, the reactive astrogliosis marker CHI3L1, the synaptic protein NPTX1, and the cerebrovascular markers PGF, PDGFRB, and VEFGA; all previously linked to AD but only reliably measured in cerebrospinal fluid. SQSTM1, the autophagosome cargo protein, exhibited a significant association with neurodegeneration status after adjusting age, sex, and *APOE* ε4 genotype.

**Conclusions:** Together, our results demonstrate the feasibility and potential of immunoassay-based multiplexing to provide a comprehensive view of AD-associated proteomic changes. Further validation of the identified inflammation, synaptic, and vascular markers will be important for establishing disease state markers in asymptomatic AD.

## Background

The draft revision of the amyloid/tau/neurodegeneration (AT(N)) research framework emphasizes the multifaceted nature of Alzheimer’s disease (AD), involving diverse brain pathologies and physiological processes [1]. In addition to A (β-amyloid deposition), T (pathologic tau), and N (neurodegeneration) categories, which were included in the 2018 update [2], the proposed upcoming revision recommends biomarker assessments for inflammation (I) as well as mixed pathologies such as vascular (V) pathology and synucleinopathy (S). Furthermore, alteration of synapses can occur early in the AD continuum, even before overt neurodegeneration, making the examination of synaptic markers important in preclinical AD [3–5]. This new framework necessitates a diverse set of biomarkers for more accurate diagnosis, prognosis, clinical management, and development/evaluation of therapies. Analyses of multiple biomarkers integrated into a single test can enhance efficiency, reduce analytical errors, and save on specimen volume. However, multi-analyte assays that provide concurrent information on A, T, and N processes are lacking, let alone those that concomitantly include I, V, and S biomarkers. In fact, glial fibrillary acidic protein (GFAP) is the only marker listed under I, while V has no entry in terms of biofluid biomarkers recommended in the draft revision of the research and diagnostic framework [1].

Previous analyses of cerebrospinal fluid (CSF) implicated associations of several inflammatory, vascular, and synaptic function proteins with amyloid-beta (Aβ) and tau pathologies in AD. Regarding neuroinflammation, the astrocytic protein chitinase-3 like-protein-1 (CHI3L1), also known as YKL-40, has been shown to associate preferentially with tau pathology, while GFAP, a different astrocytic protein, was more involved with Aβ plaque pathology [6–9]. CSF levels of soluble TREM2, a transmembrane receptor protein predominantly expressed by microglia cells, were increased in AD and associated with tau-dependent neurodegeneration and cognitive decline [10–13]. Levels of TREM1, another microglial transmembrane protein, were also shown to increase in AD dementia compared with cognitively unimpaired controls and those with mild cognitive impairment (MCI) [14]. In addition, multiple interleukins (ILs) in CSF were associated with Aβ and tau pathology, as well as cognitive decline [15–20]. Similarly, several CSF markers of cerebrovascular integrity, such as sPDGFRB (soluble platelet-derived growth factor receptor β), ICAM1, VCAM1, and VEGFs, synaptic markers such as NPTX and NRGN, have been linked to AD and cognitive decline [18, 21–28]. High α-synuclein seed amplification assay positivity has been found in AD and is associated with atypical clinical manifestation [29].

A major challenge in the AD biomarker field is the difficulty in accurately measuring the aforementioned neuroinflammation, cerebrovascular, and synaptic protein markers in blood samples to give reliable performances as shown for their CSF counterparts. The development of blood-based assays for these biomarkers has been greatly impeded by several factors, including interference from the extremely complex blood proteome, low abundance of the target analytes, and signal attenuation by unwanted signal from peripheral sources [30, 31]. For example, assays for synaptic markers including NRGN give good analytical signals in plasma but without the corresponding good biomarker performance as shown in CSF [32, 33].

Recently, a highly multiplexed immunoassay capable of measuring classical AT(N) biomarkers alongside multiple I, V, and S biomarkers in plasma has been described [34]. Known as the NULISAseq CNS disease panel, this assay employs an innovative automated technology called NUcleic acid-Linked Immuno-Sandwich Assay (NULISA). Coupling NULISA with next-generation sequencing readout (NULISAseq) allows detection of hundreds of proteins with attomolar sensitivity and an ultra-broad dynamic range [34]. The NULISAseq CNS panel consists of ∼120 protein targets covering the eight pathological hallmarks that define neurodegenerative diseases: namely, pathological protein aggregation, synaptic and neuronal network dysfunction, aberrant proteostasis, cytoskeletal abnormalities, altered energy homeostasis, DNA and RNA defects, inflammation, and neuronal cell death [35].

This study had a three-fold aim. Our first aim was to evaluate the technical performance of the NULISAseq CNS disease panel. For classical AT(N) biomarkers (e.g., p-tau181, p-tau217, p-tau231, Aβ40, Aβ42, GFAP, and neurofilament light chain [NEFL]), we compared the NULISA results with those obtained, in mostly single-plex formats, on the widely used Quanterix Single molecule array (Simoa) platform. The second aim was to examine the diagnostic accuracies and longitudinal profiles of blood-based NULISAseq targets against neuroimaging measures of A, T and N in a population-based cohort of mostly cognitively normal older adults. Thirdly, we aimed to identify novel plasma I, V, and synaptic markers associated with Aβ positron emission tomography (PET), tau PET and magnetic resonance imaging (MRI)-based neurodegeneration measures in the same cohort.

## Methods

### Participants

The Monongahela Youghiogheny Healthy Aging Team-Neuroimaging (MYHAT-NI) is a sub-cohort of the parent MYHAT study, a population-based prospective study of cognitively normal older adults designed to characterize the prevalence of MCI in older adults with a low socioeconomic status in selected Rust Belt regions in southwestern Pennsylvania [36–38]. The MYHAT study recruited participants aged 65 and older via age-stratified random sampling from publicly available voter registration lists. The MYHAT-NI study recruited a subset of MYHAT participants with a Clinical Dementia Rating (CDR) sum-of-box score [39] of < 1.0 for neuroimaging assessments to investigate the distribution and functional correlates of AD pathologies. For this reason, all MYHAT-NI participants had normal or only very mildly impaired cognition at the time of enrollment which started in 2017. The only exclusion criterion was a contraindication to neuroimaging. The study had two visits: baseline and approximately two-year follow-up. Sociodemographic information was collected at the baseline visit. Blood collection, neurophysiological assessment, and neuroimaging, including [^11^C] Pittsburgh Compound B (PiB) positron emission tomography (PET) imaging of Aβ plaques, [^18^F]AV-1451 PET imaging of tau pathology, and structural MRI for neurodegeneration, were performed at both baseline and the follow-up visits. Detailed study designs for MYHAT and MYHAT-NI, including subject recruitment strategies, multi-domain cognitive assessments, neuroimaging, and data processing, can be found in previous publications [36, 38]. *APOE* genotyping was determined as previously described [40].

We classified participants’ A, T, and N status according to [^11^C] PiB PET, [^18^F]AV-1451 PET, and MRI scans for cortical thickness, respectively. The A status was based on a global [^11^C] PiB standardized uptake value ratio (SUVR) computed by volume-weighted averaging of nine composite regional outcomes (anterior cingulate, posterior cingulate, insula, superior frontal cortex, orbitofrontal cortex, lateral temporal cortex, parietal, precuneus, and ventral striatum) [41]. Participants were classified as A+ or A-based on a pre-defined cutoff, with >1.346 as A+ [42, 43]. For T status, a composite SUVR was computed for each [^18^F]AV-1451 PET by normalizing composite Braak regional values ((Braak I - VI) to FreeSurfer cerebellar gray matter activity [44, 45]. Participants with SUVR > 1.18 were considered T+, <=1.18 as T- [46]. N status was based on an AD-signature composite cortical thickness index derived from a surface-area weighted average of the mean cortical thickness of four FreeSurfer regions of interest (ROIs) – entorhinal, inferior temporal, middle temporal, and fusiform – that are most predictive of AD-specific diagnosis and pathology, with < 2.7 as N+ [42, 47]. The MYHAT-NI study was approved by the University of Pittsburgh Institutional Review Board (STUDY19020264).

### NULISAseq assay procedures and data processing

Plasma samples were thawed and centrifuged at 10,000xg for 10 min to remove particulates. The supernatants were then analyzed using the NULISAseq CNS disease panel, an innovative proprietary proteomic platform, on an Alamar ARGO^TM^ prototype system, as previously described [34]. In brief, samples were incubated with a cocktail of paired capture and detection antibodies for the included target protein biomarkers and the internal control (IC). The capture antibodies were conjugated with partially double-stranded DNA containing a poly-A tail and a target-specific barcode, while detection antibodies were conjugated with another partially double-stranded DNA containing a biotin group and a matching target-specific barcode. After incubation, the mixtures underwent magnetic bead-based capture, wash, release, recapture, and second round of wash processes to purify the formed immunocomplexes. A ligation mix, including T4 DNA ligase and a specific DNA ligator sequence, was utilized to ligate the proximal ends of DNA attached to the paired antibodies, generating DNA reporter molecules containing unique target and sample-specific barcodes. The reporter DNA levels were then quantified by Next-Generation Sequencing (NGS). The plasma samples were randomized in two plates for the assay. Three assay controls were run side-by-side with samples for each plate, including the sample control (2 replicates/plate), the inter-plate control (3 replicates/plate), and the negative control (2-3 replicates/plate).

Data normalization was performed to remove potential unwanted technical variation. First, IC-based normalization was done by dividing the target counts for each sample well by that well’s IC counts. IPC normalization was achieved by dividing IC-normalized counts by target-specific medians of the IPC (pooled plasma) sample replicates on that plate. Finally, the data was rescaled and log2-transformed to give a more normal distribution for subsequent statistical analyses. These values are hereafter referred to as NULISA Protein Quantification (NPQ) units. The fold change difference between two groups were calculated as 2^(difference^ ^in^ ^NPQ)^. The plate-specific limit of detection (LOD) was calculated for each target assay by taking the mean plus three times the standard deviation of the unlogged normalized counts for the negative control samples on the plate. LODs were then rescaled and log2-transformed as above.

### Procedures for Simoa assays

Simoa assays were performed on an HD-X instrument (Quanterix, Billerica, MA, USA). Prior to the measurements, plasma samples were thawed at room temperature and centrifuged at 4000xg for 10 min to remove particulates. Plasma NEFL, GFAP, Aβ42 and Aβ40 were measured with the Neurology 4-Plex E (#103670), p-tau181 with the p-tau181 V2 Advantage kit (#103714), and p-tau217 with the ALZpath Simoa® p-Tau 217 V2 Assay Kit (#104371). Quality control (QC) samples of 2-3 different concentrations for each assay were analyzed at the start and the end of each run to assess the reproducibility of each assay. The average within-run coefficient of variations (CVs) of the QC samples were 3.7% for p-tau217, 6.6% for p-tau181, 14.3 for NEFL, 9.9% for GFAP, 8.9% for Aβ42, and 9.5% for Aβ40. The average between-run CVs were 11.4% for p-tau217, 11.7% for p-tau181, 18.3% for NEFL, 17.8% for GFAP, 13.0% for Aβ42, and 14.6% for Aβ40.

### Statistical analysis

All analyses were conducted using MATLAB (version R2021b) or R statistical software version 4.2.1 (R Foundation for Statistical Computing, Vienna, Austria; http://www.r-project.org/). In general, we utilized the Wilcoxon rank-sum test for two-group comparisons and the Kruskal-Wallis test for comparisons involving more than two groups. Spearman’s rank correlation was used to measure the strength and direction of association between two continuous variables. For demographic characteristics, continuous variables were presented as mean and standard deviation, while categorical variables were reported as counts. Wilcoxon rank-sum tests and Fisher’s exact tests were employed to assess the significance of differences between A+ and A-participants for continuous and categorical variables, respectively. Linear mixed models were utilized to assess the association of common AD risk factors, including age, sex, and *APOE* ε4 carrier status, with biomarkers.

The following statistical tests were utilized to explore cross-sectional associations between biomarkers and brain Aβ and tau pathologies: (1) Wilcoxon rank-sum tests for the univariate significance for the associations between NPQs and dichotomous pathology variables (e.g., A-vs. A+), without adjusting for risk factors; (2) Spearman’s rank correlation was used to measure the strength and direction of the associations between NPQs and continuous variables (e.g., Aβ PET SUVR); (3) linear mixed models (random intercepts) with biomarker NPQs as the dependent variable, visit-specific Aβ PET status as the independent variables, as well as common risk factors (such as age, sex and *APOE* ε4 carrier status) were used to determine the overall risk factor-adjusted significance combining samples from both visits. False discovery rate corresponding to cutoff p-values were calculated according to the procedure described by Yoav Benjamini and Yosef Hochberg in 1995 [48]. An arbitrary *p*-value of 0.005 was used as the significance cutoff, which corresponded to 3 to 10% FDR depending on the comparisons. Receiver operating characteristic (ROC) curves and the area under curve (AUC) were calculated using the MATLAB *perfcurve* function, based on scores predicted from generalized linear regression models fitted using the MATLAB *fitglm* function. Confidence intervals were computed using bootstrap with 1000 replicates. DeLong test implemented in the pROC package was used to compare ROC curves [49, 50]. Web app VolcaNoseR was used to create draw the volcano plot [51].

Longitudinal analysis was limited to participants with plasma samples analyzed at both visits. We calculated the yearly percentage of change for biomarker NPQs and continuous AD pathology variables using this formula: 100 * ([Follow up – Baseline]/[Baseline]) /Δ Time in years. Wilcoxon rank-sum tests were then used for two-group comparisons, and Spearman’s rank correlation was used to measure the association between yearly plasma biomarker changes and the yearly AD pathology change. Due to the relatively short duration between the two visits, we did not expect drastic changes in both blood and neuroimaging biomarker levels. Therefore, we treated the longitudinal analysis as explorative, and the original rather than FDR-adjusted *p*-values were used to determine significance.

## Results

### Cohort characteristics

This study comprised 176 plasma samples from 113 participants (average age 76.7 years at baseline, 54.0% women, and 95.0% non-Hispanic White) from the MYHAT-NI cohort (see Table 1 for demographic characteristics). Among them, 63 participants (55.8%) provided plasma samples at two visits (baseline and the 2-year visit). At baseline, 85 (75.2%) participants were classified as A-negative (A-) and 28 (24.8%) as A-positive (A+), while 42 (66.7%) and 21 (33.3%) were A- and A+, respectively at the 2-year visit. Regarding tau PET, 74 (65.4%) participants were classified as tau-negative (T-) and 39 (34.5%) as tau-positive (T+) at baseline. At the 2-year visit, 42 (66.7%) participants were T-, and 21 (33.3%) were T+. In terms of neurodegeneration according to cortical thickness, 80 (70.8%) participants were considered N- and 33 (29.2%) N+ at baseline, while at the 2-year visit, there were 42 (66.7%) N- and 20 (31.7%) N+ participants. One participant had missing N status at the 2-year visit due to poor MRI quality.

Most participants were cognitively normal at both visits. Clinical Dementia Rating (CDR)-based cognitive assessment rated 101 participants (89.3%) as cognitively normal (CDR=0) and 10 (8.9%) as mildly impaired (CDR=0.5) at baseline. At the 2-year visit, 55 participants (87.2%) were cognitively normal, and 5 (7.9%) were mildly impaired. Two and three participants missed CDR assessments at baseline and the 2-year visit, respectively. Similarly, Mini-Mental State Examination (MMSE) assessment rated 107 (94.6%) participants as cognitively normal (MMSE >=24) and 4 (3.5%) as mildly impaired (MMSE between 19 – 23) at baseline, while all participants at the 2-year visit were cognitively normal.

### Technical performance and head-to-head comparison of the NULISAseq measurements with Simoa assays

A total of 116 target assays were incorporated in the NULISAseq CNS disease panel for this study. The plasma concentration range of these targets spanned a minimum of 6 orders of magnitude according to the concentration estimated by mass spectrometry-based proteomics in the Human Protein Atlas database [52, 53]. Despite the broad dynamic ranges of the protein targets, the vast majority exhibited very high detectability, defined as the percentage of samples above the LOD, with a mean ± standard deviation (SD) detectability of 97.2% ± 13.9% (Fig. 1A). Only three targets – UCHL1, PTN, and pTDP43-409 – had detectability below 70%. The 176 plasma samples, including 113 baseline and 63 2-year visit samples, were distributed across two plates for NULISAseq measurements. Each plate included two replicates of a pooled plasma sample to assess assay reproducibility. The median intra-plate and inter-plate CVs were 4.34% (interquartile range [IQR]: 2.80%-6.04%) and 3.11% (IQR: 1.41% -5.45%), respectively, suggesting robust assay reproducibility (Fig. 1B and 1C). Only two targets – CNTN2 and NEFH – had inter-plate CVs greater than 20%, a cutoff commonly used for in vitro diagnostic assays. To assess whether the variation depended on protein abundance, we evaluated the association between the intra- or inter-plate CVs and the abundance ranks for targets (n=46) with plasma concentration data available from the Human Protein Atlas database. As depicted in Fig. 1D and 1E, both intra- and inter-plate CVs were not influenced by protein abundance, with *p*-values for Spearman rank correlations being 0.173 and 0.919, respectively.

**Fig. 1:**
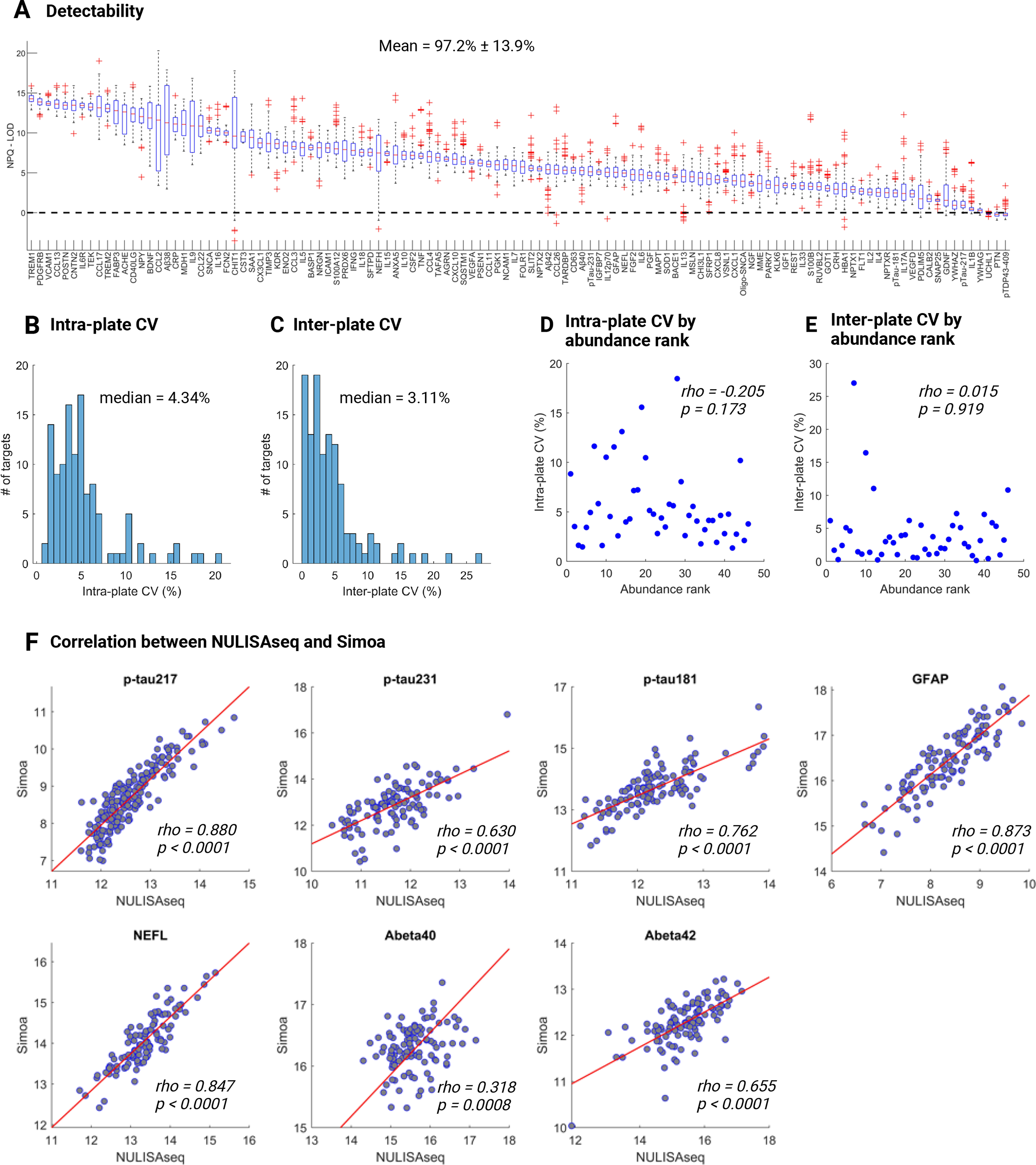
Performance of the NULISAseq CNS disease panel. **A** Box plots illustrating the detectability of 116 targets in 176 plasma samples collected from 113 MYHAT-NI participants. The y-axis represents NPQ - LOD, where values >0 indicate detectability. On each box, the central mark indicates the median, and the bottom and top edges of the box indicate the 25th and 75th percentiles, respectively. The whiskers extend to the most extreme data points not considered outliers, and the outliers are plotted individually using the ’+’ marker symbol. Data points were considered outliers if they were greater than q3 + 1.5 × (q3 – q1) or less than q1 – 1.5 × (q3 – q1), where q1 and q3 are the 25th and 75th percentiles of the sample data. **B-C** Histogram distribution of intra-plate (**B**) and inter-plate (**C**) coefficient of variations (CVs). **D-E** Scatterplot distributions between abundance rank and intra-plate (**D**) or inter-plate (**E**) CVs. Intra- and inter-plate CVs were calculated based on results of a pooled plasma sample (SC), measured in duplicate each in two different plates. Abundance rank was based on the mass spectrometry-estimated protein abundance in the Human Protein Atlas (downloaded on 12/24/2023). (**F)** Scatterplot distributions illustrating the correlation of protein levels measured using NULISAseq and Simoa method. *Rho* and *p* values were determined using Spearman rank-based correlation. Purple lines indicated the least square regression lines. NPQ, NULISA Protein Quantification; LOD, limit of detection.

We next examined the correlation between NULISAseq measurements and Simoa measurements of selected biomarkers. These included p-tau217, p-tau231, p-tau181, GFAP, NEFL, Aβ40, and Aβ42. Notably, strong correlations were observed in all pairwise comparisons, with Spearman rank correlation coefficient (*rho*) values spanning from 0.318 to 0.880 (Fig. 1F). P-tau217, GFAP, and NEFL demonstrated the strongest between-platform correlation, with *rho* of 0.880, 0.873, and 0.847, respectively.

To compare the diagnostic accuracies of the two measurements in detecting Aβ PET positivity, we calculated the ROC AUCs using logistic regression models in the baseline samples (Table 2). NULISAseq demonstrated comparable performance to Simoa for all seven biomarkers, irrespective of whether common risk factors (age, *APOE* ε4 carrier status, and sex) were included in the models. For example, at baseline, plasma p-tau217 had AUCs of 0.905 (95% CI: 0.841-0.969) on NULISA and 0.880 (95% CI: 0.800-0.959) on Simoa. When accounting for risk factors, these AUCs increased to 0.930 (95% CI: 0.878-0.980) and 0.925 (95% CI: 0.874-0.977). The DeLong test showed no significant difference between the AUCs. These findings suggest that despite its highly multiplexed nature, the NULISAseq platform performs equivalently as Simoa for quantifying these biomarkers.

### Association of NULISAseq targets with PET measure of amyloid pathology (A)

#### Cross-sectional association

Several NULISAseq targets showed significant association with the common AD risk factors age, sex, and *APOE* ε4 carrier status (Additional file 1: Figure S1). To account for the potential confounding effect of these risk factors, we utilized linear mixed models, as described in the “Materials and methods” section, to evaluate the adjusted significance for the cross-sectional association between NULISAseq targets and neuroimaging biomarkers.

A total of 16 targets showed significant association with Aβ pathology, as determined by Aβ PET, according to *p*-value < 0.005, corresponding to approximately 8% FDR (Fig. 2A). Fig. 2B illustrates boxplot distributions of significant targets at baseline and the 2-year visit. NULISAseq plasma p-tau217 demonstrated superior diagnostic accuracy in detecting Aβ PET positivity. As stated above, plasma p-tau217 achieved AUCs of 0.905 (95% CI: 0.841-0.969) and 0.922 (95% CI: 0.825-0.972) in the baseline and 2-year visit, respectively, when utilized as the sole predictor. Incorporating common AD risk factors – age, sex, and *APOE* ε4 carrier status – raised the AUCs to 0.930 (95% CI: 0.878-0.983) and 0.938 (95% CI: 0.856-0.979), respectively. On average, A+ participants exhibited an 82.8% elevation in plasma p-tau217 levels compared to A-controls. NULISAseq p-tau231 also exhibited significant association. However, the AUCs and fold increases were inferior to plasma p-tau217. The AUCs were 0.718 (95% CI: 0.615-0.822) in the baseline and 0.698 (95% CI: 0.556-0.821) in the 2-year cohorts based on biomarker-only models, which increased to 0.808 (95% CI: 0.717-0.899) and 0.794 (95% CI: 0.652-0.888) respectively, with the inclusion of common risk factors. An overall 30.7% increase was observed comparing p-tau231 levels in A+ participants to those in A-controls. Moreover, GFAP showed high univariate association prior to adjusting for common risk factors, with Wilcoxon rank-sum *p*-values of 0.0002 for the baseline and 0.006 for the 2-year cohort. However, GFAP showed a strong association with age and APOE ε4 carrier status (Additional file 1: Figure S1B and S1C), and its risk factor-adjusted significance weakened to a *p*-value of 0.016. The fold increase of GFAP in A+ vs. A-participants was 45.7%. It distinguished A+ from A-participants with AUCs of 0.732 (95% CI: 0.640-0.825) and 0.715 (95% CI: 0.569-0.830) in the baseline and 2-year cohorts based on biomarker-only models, and 0.808 (95% CI: 0.717-0.899) and 0.815 (95% CI: 0.668-0.926), respectively, after adjusting for common risk factors.

**Fig. 2:**
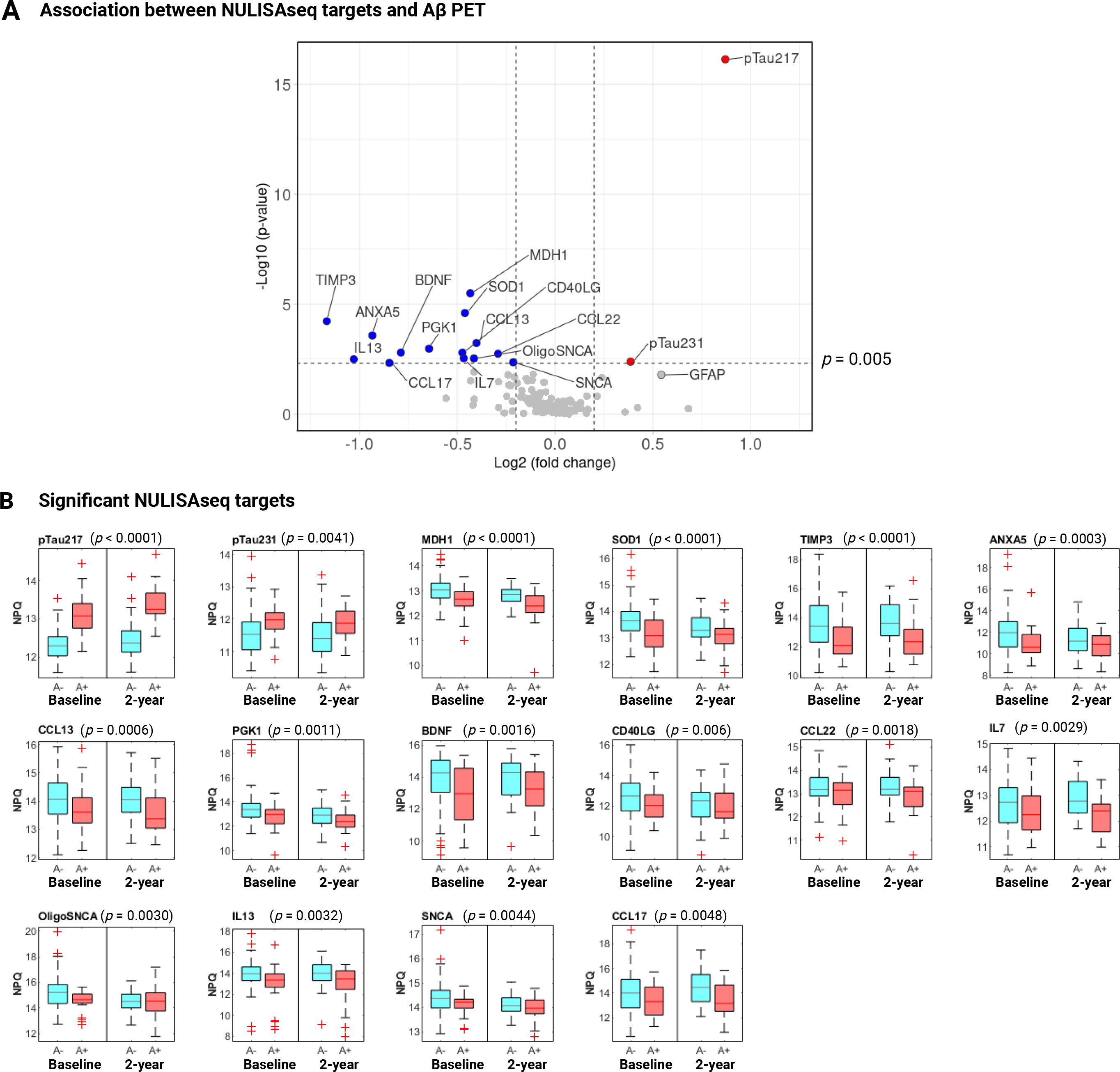
Cross-sectional association of NULISAseq targets with amyloid pathology (A). **A** Volcano plot of -log10(p-value) versus log2(fold change) comparing biomarker abundances (NPQ) in samples from A+ participants (n=49) vs. A-controls (n=127). Significant targets are shown in red (higher in A+) or blue (lower in A+) circles. Grey circles represent non-significant targets. **B** Boxplot distributions of significant NULISAseq targets, separated by A status and visit. P-values on top of the boxplots were for the whole data combining both visits and were determined using linear mixed models (random intercepts) with NPQs as the dependent variable, visit-specific A status as the independent variables, adjusting for covariates age, sex, and APOE ε4 carrier status. Significance determination was based on p-value < 0.005, corresponding to ∼8% FDR.

Contrarily, the other targets that showed significant associations with Aβ pathology showed decreased protein levels in the A+ versus A-participants (Fig. 2A and 2B). Metalloproteinase inhibitor 3 (TIMP3), a metalloprotease inhibitor involved in regulating proteostasis [54, 55], exhibited the most substantial decrease in protein levels, with a 60%-fold decrease in A+ vs. A-individuals. TIMP3 distinguished A+ and A-participants with AUCs of 0.711 (95%CI: 0.596-0.814) and 0.739 (95%CI: 0.574-0.859) for the baseline and 2-year visit cohorts when used as the sole predictor. The inclusion of common risk factors improved the AUCs to 0.850 (95% CI: 0.718-0.923) and 0.882 (95% CI: 0.757-0.946). Malate dehydrogenase subunit 1, MDH1, also emerged as one of the top significant proteins, with the risk factor-adjusted *p*-value < 0.0001, and decreased at an average of 26% in A+ participants. BDNF, a neurotrophic factor with pivotal roles in regulating synaptic plasticity and neuronal survival, showed an overall 42% reduction in A+ participants.

Six cytokines—IL7, IL13, CD40LG, CCL13, CCL17, and CCL22—were significantly associated with Aβ pathology, supporting the involvement of immune response and inflammation in AD. Additional significant targets included SOD1, PGK1, ANXA5, and NULISAseq targets for soluble α-synuclein (SNCA) and oligomeric α-synuclein (OligoSNCA).

#### Longitudinal association

We then investigated the longitudinal relationship between NULISAseq targets and Aβ pathology. Three cytokines – FGF2, IL4, and IL9 – exhibited Aβ PET-dependent yearly percentage changes, with Wilcoxon rank-sum test *p*-values of 0.02, 0.04, and 0.04, respectively (Fig. 3A). The median yearly percentage changes were 9.1%, 1.5%, and 15.4% in A+ individuals, compared to -12.8%, -9.5%, and 0.4% in A-participants, respectively, for FGF2, IL4, and IL9. This means that while the cytokine levels increased over time in A+ individuals, large decreases were recorded for A-participants.

**Fig. 3:**
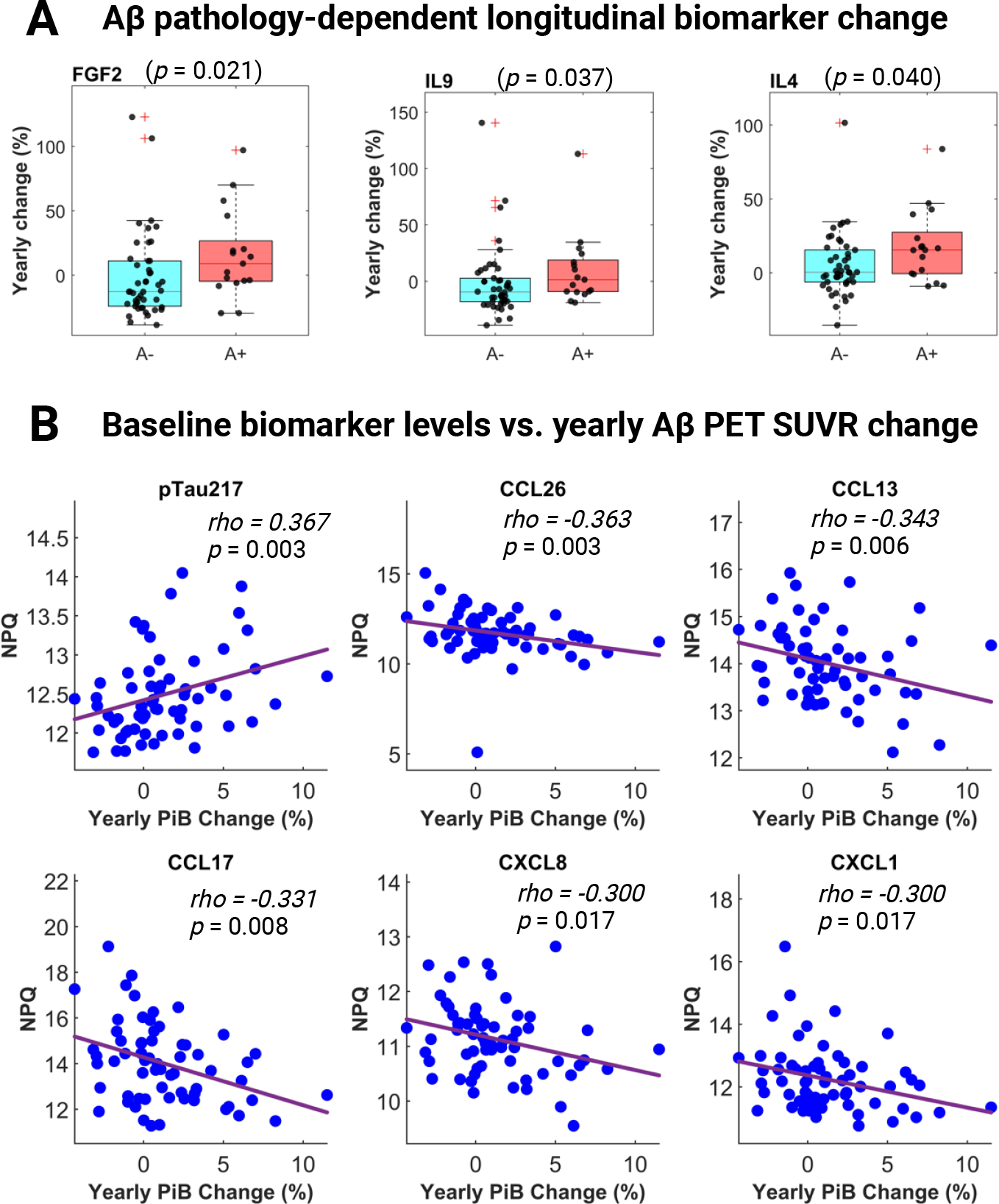
Longitudinal association between NULISAseq targets and amyloid pathology (A). **A** Boxplots illustrating the distribution of yearly biomarker abundance change by A status. P-values were based on two-sided Wilcoxon rank sum tests. **B** Scatterplots for the correlation between yearly longitudinal Aβ PET SUVR change and baseline biomarker levels. The strength of the correlation was assessed based on Spearman’s ranks. Purple lines indicated the least square regression lines.

Next, we explored the influence of baseline biomarker levels on the progression of Aβ pathology, defined as the yearly percentage of change in Aβ PET SUVR. Apart from p-tau217, five chemokines – CCL26, CCL17, CCL13, CXCL1, and CXCL8 – demonstrated significant associations (Fig. 3B). Higher baseline levels of p-tau217 were associated with more robust increases in Aβ PET SUVR, with Spearman *rho* of 0.367 (*p* = 0.003). On the contrary, elevated baseline levels of all five chemokines were linked with a smaller Aβ PET SUVR increase, with *rho* of -0.363 (*p* = 0.004) for CCL26, -0.343 (*p* = 0.006) for CCL13, -0.331 (*p* = 0.008) for CCL17, -0.300 (*p* = 0.017) for CXCL18, and -0.300 (*p* = 0.017) for CXCL1. Given that a slower increase in Aβ PET SUVR changes is likely indicative of a more favorable prognostic outcome, these findings suggest that higher levels of these chemokines may confer a protective role in recruiting immune cells to attenuate the accumulation of Aβ plaques. Consistent with this, all five chemokines were lower in abundance in A+ participants, albeit only CCL13 and CCL17 passed the significance cutoff.

We further tested the association of plasma biomarker longitudinal changes with Aβ PET SUVR changes. Seven NULISAseq targets, namely IL5, p-tau217, Aβ38, PGF, CCL2, IL4, and VEGFD, showed strong correlations (Additional file 1: Figure S2). The changes of all seven targets were positively correlated with Aβ PET SUVR changes, suggesting that upward changes of these targets over time might be correlated with more severe Aβ pathology.

### Association of NULISAseq targets with tau pathology (T)

#### Cross-sectional association

Five NULISAseq targets displayed significant associations with tau PET positivity according to *p*-value cutoff of 0.005, corresponding to FDR of 9%, after adjusting for age, sex and *APOE* ε4 carrier status (Fig. 4A). The three top significant targets were p-tau species, namely p-tau231 (*p* = 0.0004), p-tau217 (*p* = 0.0005), and p-tau181 (*p* = 0.003). SFRP1, a Wnt signaling modulator [56], and YWHAG, a member of the 14-3-3 family proteins, were the other significant targets, with *p*-values of 0.003 and 0.004, respectively. All except SFRP1 were increased in T+ participants. Average fold increases of 29%, 36%, 20%, and 5% in T+ participants compared with T-controls were observed for p-tau231, p-tau217, p-tau181, and YWHAG, respectively. SFRP1, on the other hand, was decreased at an average of 27%. Among these five targets, p-tau217 had the highest diagnostic accuracy in detecting abnormal tau pathology, with AUCs of 0.652 (95% CI: 0.518-0.765) for the baseline cohort and 0.797 (95% CI: 0.660-0.888) for the 2-year cohort. This was followed by p-tau231, which had AUCs of 0.651 (95%CI: 0.522-0.759) and 0.705 (95% CI: 0.560-0.816), respectively. The inclusion of age, sex, and *APOE* ε4 carrier status only slightly improved the AUCs (Additional file 1: Figure S3). Both p-tau217 and p-tau231 showed better diagnostic accuracies in the 2-year cohort, consistent with the expectation that tau pathology worsens over time and that agreement between the plasma and neuroimaging biomarkers improves with disease progression.

**Fig. 4:**
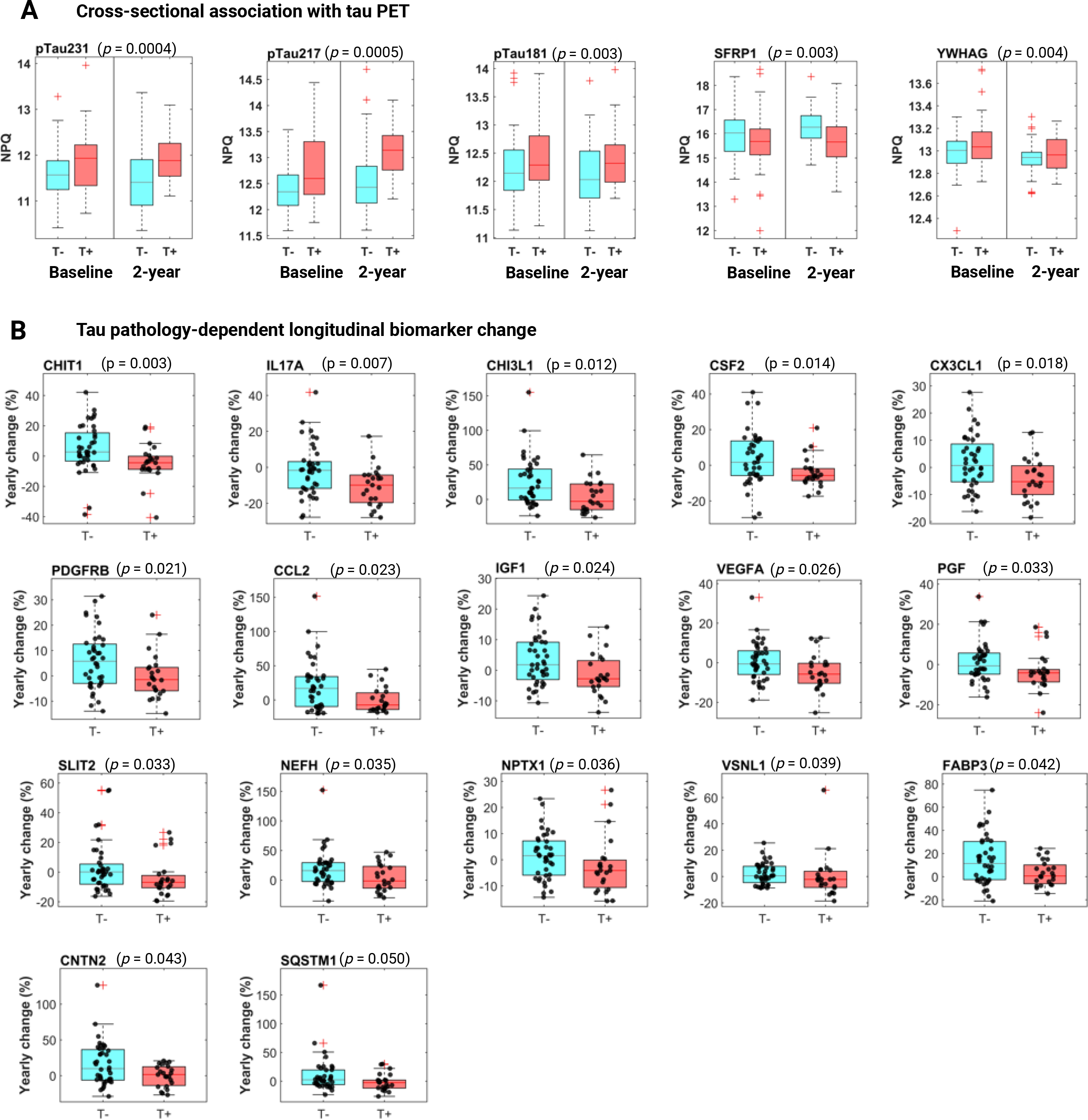
Association of NULISAseq targets with tau pathology (T). **A** Boxplots of NULISAseq targets with significant cross-sectional associations with T status, separated by T status and visit. P-values on top of the boxplots were for the whole data combining both visits and were determined using linear mixed models (random intercepts) with NPQs as the dependent variable, visit-specific T status as the independent variables, adjusting for covariates age, sex and APOE ε4 carrier status. Significance determination was based on p-value < 0.005, corresponding to ∼9% FDR. **B** Boxplots illustrating the distribution of yearly biomarker abundance change by T status. P-values were based on two-sided Wilcoxon rank sum tests.

As indicated in Table 1, there was a higher proportion of A+ individuals among those T+. To test whether the Aβ PET status contributed to the observed association between these significant targets and tau PET status, we evaluated the association by including Aβ PET status in the linear mixed models. All remained significant except for p-tau217, which was trending towards significance (*p* = 0.068) after adjusting for the effect of Aβ PET status. The *p*-values after adjusting for Aβ PET status were 0.005, 0.003, 0.005, and 0.001 for p-tau231, p-tau181, SFRP1, and YWHAG, respectively.

#### Longitudinal association

A total of 17 targets displayed significant tau pathology-dependent longitudinal changes according to the Wilcoxon rank-sum *p*-value < 0.05 (Fig. 4B). Chitotriosidase-1 (CHIT1), a known indicator of microglial activation [57, 58], emerged as the top significant target (*p* = 0.003). Its levels showed slight increases in T-participants, with a median yearly change of 2.7%. T+ participants, on the contrary, exhibited a median yearly decrease of 4.4%. Similarly, CHI3L1 (YKL-40), a biomarker for reactive astrogliosis, also exhibited an increase (median yearly change of 16.3%) in T-participants but a decrease in T+ individuals (median yearly change of -2.8%). These observations suggest that Aβ pathology may trigger early activation of the brain’s immune system to mitigate damage, but this response may plateau or decrease as more downstream pathology, such as tau pathology, becomes apparent. Alternatively, lower glial activation in response to amyloid and tau pathology may reflect the resilience of pathologically burdened but cognitively preserved individuals [59, 60].

PGF, PDGFRB, and VEFGA, important players in maintaining cerebrovascular integrity, also showed tau pathology-dependent longitudinal changes. All three exhibited a declining trend over time in T+ participants (median yearly change: PGF, -4.1%; PDGFRB, -1.5%; VEGFA, -5.7%), contrasting with either stable or increased levels observed in T-individuals (median yearly change: PGF, -0.6%; PDGFRB, 5.8%; VEFGA, -0.6%). These findings suggest that tau pathology may be linked to the deterioration of vascular structure. NPTX1, a biomarker of excitatory synaptic pathology, similarly displayed a decreasing trend in T+ participants (median - 4.1%/year), in contrast to a slight upward change in T-controls (median 1.6%/year). Additional targets with tau pathology-dependent longitudinal changes included 4 cytokines (CCL2, CSF2, IL17A, and CX3CL1), proteins involved in synaptic and neuronal dysfunction (VSNL1, CNTN2, and FABP3), IGF1, SLIT2, NEFH, and SQSTM1.

Baseline levels of three NULISAseq targets, IL12p70, IFNG, and RUVBL2, were significantly associated with tau PET changes between the two visits (Additional file 1: Figure S4A). High baseline levels of IL12p70 and IFNG, two presumptive pro-inflammatory cytokines, were associated with faster progression of tau pathology, as determined by a more pronounced increase in tau PET composite, with *rho* of 0.288 (*p* = 0.022) and 0.269 (*p* = 0.033), respectively. The opposite relationship was recorded for RUVBL2, an AAA-type ATPase involved in regulating pro-inflammatory response. A higher baseline level of RUVBL2 was associated with a smaller increase in tau pathology, with a rho of -0.253 (*p* = 0.046).

The longitudinal change of seven NULISAseq targets correlated significantly with tau PET SUVR change (Additional file 1: Figure S4B). Interestingly, the list included three tau targets, all of which showed positive associations, namely MAPT (t-tau; *rho* = 0.348; *p* = 0.005), p-tau217 (*rho* = 0.293; *p* = 0.020) and p-tau181 (*rho* = 0.251; *p* = 0.047). Other targets on the list included SOD1 (*rho* = 0.359; *p* = 0.004), IL6R (*rho* = 0.292; *p* = 0.020), CSF2 (*rho* = 0.280; *p* = 0.027), and CD40LG (*rho* = -0.262; *p* = 0.038).

### Association of NULISAseq targets with neurodegeneration (N)

#### Cross-sectional association

Twenty NULISAseq targets exhibited significant associations with N status when assessed using univariate analysis with dichotomous outcomes (N-vs. N+) or Spearman correlations with MRI-determined cortical thickness (*p*-value < 0.005, corresponding to ∼ 5% FDR) without adjusting for the effects of common risk factors. The list of significant targets included NEFL, a classical biomarker for neurodegeneration, 8 cytokines (IL2, IL6, IL10, IL16, TNF, CCL3, CXCL10, and TAFA5), proteins previously linked to synaptic and neuronal network defects (CALB2, FABP3, and REST), proteins involved in regulating proteostasis (PSEN1, and SQSTM1), proteins involved in acute-phase response (CRP, SAA1 and SAA2), and ICAM1 and VEGFA, both critical for maintaining cerebrovascular integrity. As depicted in the heatmap in Fig. 5A, these targets exhibited a consistent trend in both the baseline and 2-year visit samples, with all targets upregulated in N+ individuals compared to N-controls. The repressor element-1 silencing transcription factor (REST), a zinc finger transcription factor with potential neuroprotective function [61], was one of the top significant targets, with *p*-values of 0.004 and 0.0008 and AUCs of 0.671 (95% CI 0.561-0.769) and 0.766 (95% CI 0.600-0.883) for the baseline and 2-year visit, respectively (Fig. 5B). NEFL showed a strong association in the baseline samples (*p* = 0.003) but not in the 2-year visit samples (*p* = 0.126) (Fig. 5B).

**Fig. 5:**
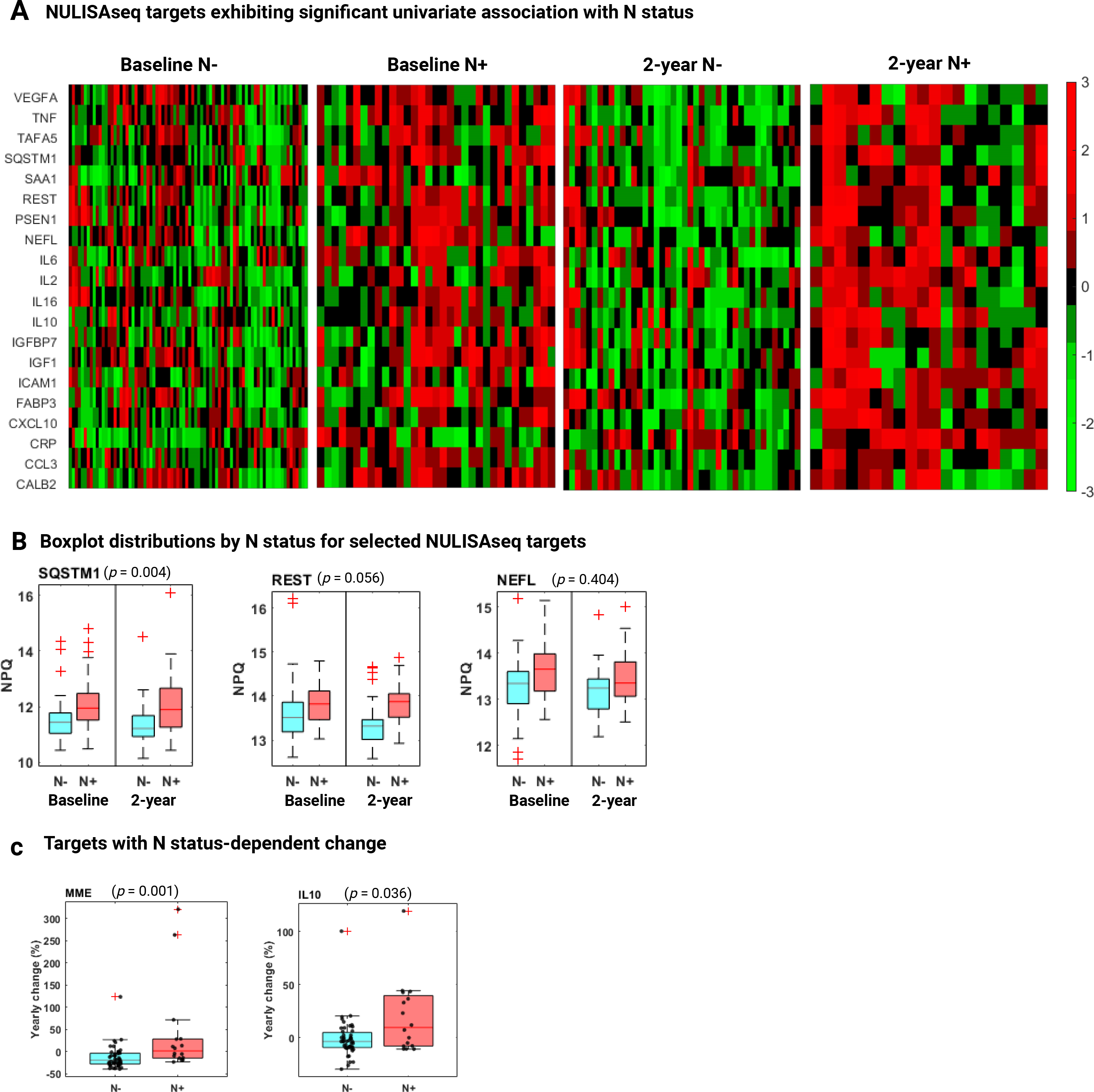
Association of NULISAseq targets with neurodegeneration (N). **A** Heatmaps illustrating the abundance levels of NULISAseq with significant univariate association with N status (unadjusted for covariates). The NPQ values were standardized for each protein target using z-scores. **B** Boxplots of selected NULISAseq targets, separated by N status and visit. P-values on top of the boxplots were for the whole data combining both visits and were determined using linear mixed models (random intercepts) with NPQs as the dependent variable, visit-specific N status as the independent variables, adjusting for covariates age, sex, and APOE ε4 carrier status. **C** Boxplots illustrating the distribution of yearly biomarker abundance change by N status. P-values were based on two-sided Wilcoxon rank sum tests.

However, after adjusting for age, sex, and *APOE* ε4 carrier status, the associations weakened for most of these targets, with SQSTM1 being the only target retaining a *p*-value < 0.005. SQSTM1 exhibited strong association with N status in both baseline and 2-year cohorts, with Wilcoxon rank-sum *p*-values of 0.0001 and 0.006, respectively (Fig. 5B). It distinguished N+ from N-participants with accuracies of 0.729 (95% CI 0.606-0.829) at baseline and 0.713 (95% CI 0.548 -0.854) at the 2-year visit (Fig. 5B). The inclusion of common risk factors (age, sex, and *APOE* ε4 carrier status) improved the accuracies to 0.776 (95% CI 0.674-0.860) and 0.848 (95% CI 0.703 -0.933).

#### Longitudinal association

MME and IL10 demonstrated neurodegeneration-dependent abundance changes, with increases in N+ participants (median change/year: MME, 1.7%; IL10, 9.4%) and decreases in N-individuals (median change/year: MME, 19.2%; IL10, 3.8%) (Fig. 5C).

Baseline levels of five NULISAseq targets – KLK6, CCL11, ARGN, TNF, and PGK1 – were significantly associated with cortical thickness change, all showing positive correlations, suggesting higher levels of these targets may be linked with a slower rate of neurodegeneration (Additional file 1: Figure S5A). The longitudinal change of five targets – PTN, YWHAZ, GOT1, NRGN, Aβ42, and SNCA (oligomer) – was significantly correlated with cortical thickness change (Additional file 1: Figure S5B). Among them, Aβ42 exhibited a positive correlation with cortical thickness change, with *rho* of 0.264 (*p* = 0.037), suggesting that a decrease in Aβ42 levels is associated with more severe neurodegeneration, i.e., a decrease in cortical thickness. Conversely, changes in the other four targets were negatively associated with cortical thickness change, with *rho* of -0.335 (*p* = 0.008), -0.291 (*p* = 0.021), -0.290 (*p* = 0.022), -0.284 (*p* = 0.025), for PTN, YWHAZ, GOT1, and NRGN, respectively.

## Discussion

In this study, we have demonstrated the feasibility of concurrent immunoassay-based analysis of 116 protein markers in blood to provide diagnostic and prognostic information in preclinical AD. Our results identified several novel inflammation, synaptic, and vascular markers in blood significantly associated with brain Aβ, tau, and neurodegeneration burden at baseline and at the two-year follow-up. These were not limited to markers such as p-tau217, p-tau231, p-tau181, and GFAP, the elevation of which have consistently shown strong associations with brain Aβ and/or tau load but included novel protein targets that inform about the disease state of the individual in different pathological stages across the biological AD continuum. Importantly, this is the first time several of these protein targets have shown validated technical and clinical biomarker potential in blood. These included the cerebrovascular markers ICAM1, VCAM1, PDGFRB, PGF, VEGFA, and VEGFD, the synaptic marker NPTX1, and the glial markers CHIT1 and CHI3L1 (YKL-40).

Concurrent measurement of a large number of protein analytes presents technical challenges that most available immunoassay platforms struggle to address. Problems such as reagent cross-activity and the dynamic range of target analyte abundance impede the multiplexing capacity of immunoassays. Technological breakthroughs, including antibody arrays, proximity ligation assay (PLA), proximity extension assay, microsphere bead capture technology by Luminex, and slow off-rate modified aptamer assay (SOMAscan), have enabled the simultaneous measurement of hundreds to thousands of plasma proteins [62]. However, these technologies often use single antibodies that only provide partial information about the protein targets, neglecting ubiquitous post-translational modifications that they undergo *in vivo*. The NUcleic acid-Linked Immuno-Sandwich Assay (NULISA) technology, which is built as an advancement of the PLA technique, integrates multiple mechanisms to enhance the performance of PLA, including a proprietary sequential immunocomplex capture and release mechanism for background reduction, next-generation sequencing-based signal readout, and fine-tuning the ratio of unconjugated “cold” antibodies to DNA-conjugated “hot” antibodies to mitigate sequencing reads of high-abundant proteins. This provides the proteomic platform the capability to detect hundreds of protein biomarkers with attomolar sensitivity and ultrabroad dynamic range [34]. The high detectability rate and low detection limits for the various protein targets in this study support this.

The strong correlation and comparable diagnostic accuracies in the head-to-head comparisons with Simoa assays indicate that both techniques measure equivalent pools of the protein targets available in the blood. It is worth mentioning that the exceptional correlation with the Simoa ALZPath assay could be due to the two assays using the same p-tau217 monoclonal antibody.

The high performance of plasma p-tau217, p-tau231, and GFAP to identify abnormal Aβ PET scans in this mostly cognitively normal cohort is corroborated by findings from several recent studies based on results from other analytical platforms [9, 63–67]. Importantly, we identified biomarkers with decreased levels in A+ participants, akin to plasma Aβ42 and Aβ42/40, indicating reduced availability in blood with progressive bran Aβ pathology. The decreases in TIMP3 are consistent with previously reported lower TIMP3 levels in AD patients [68]. TIMP3 also promotes brain Aβ production via inhibiting α-secretase cleavage of the amyloid precursor protein [69]. Reduction in MDH1 levels supports previous reports on the involvement of altered energy metabolism in late-onset AD [70, 71] while BDNF has been implicated in a protective role against A-induced neurotoxicity [72]. Several studies have also linked multiple inflammatory cytokines to Aβ pathology in AD cases and those resilient to AD [59, 73, 74]. FGF2 gene transfer reversed hippocampal function and cognitive decline in mouse models [75]. Similarly, beneficial effects of IL4 have been reported in animal models [75].

Plasma p-tau217, p-tau231, and p-tau181 were the leading markers to identify abnormal tau-PET scans. However, accounting for Aβ PET status in the combined A and T positivity analysis suggested that the results were partly explained by the strong association of these markers with Aβ pathology. This could mean that the tau forms containing these phosphorylation sites become available in blood in the early phases of Aβ plaque pathology. Aside from blood-based tau markers, YWHAG [76–78] and SFRP1 [79] showed strong associations with AD. The reduction in SFRP1 levels might be explained by its direct binding to Aβ plaques [80]. Importantly, the existing evidence from these markers has been built in CSF and brain tissue samples. Here, we extend these findings to blood.

The reactive astrogliosis marker CHI3L1 (YKL-40) and the microglia activation marker CHIT1 have repeatedly been shown to be associated with tau pathology; however, the evidence base has only been built using CSF samples [6, 81, 82]. The same applies to the vascular markers PGF, PDGFRB, and VEFGA, and the synaptic marker NPTX1, the biomarker potential of which have been demonstrated in CSF [21, 83–89]. Translation of these prior findings to plasma indicates that the molecular processes in AD involving these markers are reflected in the blood stream, expanding the repertoire of blood-based indicators of brain pathophysiological changes.

Our study identified several plasma biomarkers with a strong association with neurodegeneration assessed based on MRI-determined cortical thickness, including NEFL, which is a proven general marker of neuronal injury [90, 91]. Not surprisingly, several cytokines, including IL2, IL6, IL10, IL16, TNF, CCL3, CXCL10, and TAFA5, also showed significant association with neurodegeneration, reinforcing the close relationship between neuroinflammation and neurodegenerative processes [92, 93].

Two vascular proteins, ICAM1 and VEGFA, were on the significant list, consistent with the expected involvement of neurovascular dysfunction in neuroinflammation and neurodegeneration [94]. ICAM1 is a transmembrane glycoprotein expressed in multiple cell types and plays a key role in maintaining the blood brain barrier (BBB) [95]. Its expression is induced by neuroinflammation, leading to increased leukocyte transmigration across the BBB, a pivotal event in the pathogenesis of various brain diseases, including AD [96–98]. Consistent with this, our study observed elevated ICAM1 levels in participants with neurodegeneration, aligning with previous research [18, 20]. VEGFs have complex associations with neurological diseases, exhibiting neuroprotective and neuro-destructive potential [24, 25, 99]. We observed elevated VEGFA levels in N+ participants and a faster decline in VEGFA levels in T+ participants, suggesting a potential staging effect. Interestingly, a recent study showed that a low level of VEGFA, measured with assays from Meso Scale Discovery, was associated with accelerated neocortical tau accumulation in preclinical A+ participants in the Harvard Aging Brain Study [100].

Synaptic and neuronal network dysfunction, along with aberrant proteostasis, represent two of the eight pathological hallmarks of neurodegenerative diseases [35]. In alignment with this, our study found significant associations between neurodegeneration, three synaptic/network proteins (CALB2, FABP3, and REST), and two proteostatic regulators (PSEN1 and SQSTM1). Notably, among all targets with significant association with neurodegeneration, only SQSTM1 withstood corrections for age, sex, and *APOE* ε4 genotype, revealing it as a potentially novel neurodegeneration biomarker for AD. SQSTM1, a scaffold protein with a critical role in macroautophagy, has been previously linked to several neurodegenerative diseases, including AD [101–103].

IL10 and MME were the top hits with differential longitudinal change in N+ vs. N-participants. Several studies support their roles as markers of neurodegeneration status. For example, an animal model study suggested that the mechanisms of action of IL-10 as an inflammatory response might be through the activation of microglia, which leads to IL-6 activation and abnormal phosphorylation of tau [104]. MME, also known as neprilysin, is an integral membrane-bound metallopeptidase (MMP) and one of the key enzymes involved in Aβ degradation [105]. MMPs have been found to exhibit dual roles in AD pathogenesis. On the one hand, they can reduce the amount of Aβ deposits by degrading Aβ peptides [106, 107]. On the other hand, their levels can be induced by Aβ, potentially leading to brain parenchymal destruction [108, 109]. Interestingly, in addition to demonstrating significant longitudinal changes between N+ and N-participants, MME exhibited lower levels in A+ participants compared to A-controls, yet higher levels in N+ participants compared to N-, supporting its potential utility as a staging biomarker for AD.

Interestingly, while a number of cytokines (IL2, IL6, IL10, IL16, TNF, CCL3, CXCL10, and TAFA5) were increased in participants with neurodegeneration, several cytokines (IL7, IL13, CD40LG, CCL13, CCL17, and CCL22) were found to be decreased in participants with Aβ pathology. Additionally, higher baseline levels of several chemokines (CCL26, CCL17, CCL13, CXCL1, and CXCL8) were significantly associated with slower progression of Aβ pathology. Given that most participants with Aβ pathology were cognitively normal, and neurodegeneration is presumed to occur at a later stage than the early phase of Aβ pathology, our results support the biphasic roles of neuroinflammation, with protective effects in the early stages and potentially detrimental effects in the later stages. These findings are in line with the recognized multifaceted impact of neuroinflammation on AD pathogenesis [110, 111].

Strengths of this study include (i) technical validation of the new NULISA platform; (ii) direct comparison of the clinical performances of biomarkers measured using NULISA assays vs. with Simoa assays; (iii) focus on a population-based cohort to provide information closer to the real world than most clinical research-based cohorts; (iv) emphasis on predominantly cognitively normal participants with emerging pathological phenotypes, to test the sensitivity of the NULISA platform to these incipient changes; (v) availability of paired neuroimaging measures of Aβ, tau, and neurodegeneration, making it possible to identify inflammatory, vascular and synaptic markers associated with abnormal changes in different biologically defined disease stages; and (vi) repeated neuroimaging evaluations and blood collection over a two-year interval, allowing to examine biomarker changes within that timeframe. Limitations include the lack of validation in diverse cohorts.

### Conclusions

Together, this targeted proteomic study has established that results from the NULISA platform are equivalent to those from Simoa HDX. Additionally, the strong multiplexing capabilities of NULISA allowed for the evaluation of dozens of verified and putative protein biomarkers in a longitudinal preclinical AD cohort. We identified several neuroinflammation, synaptic, and vascular markers that have been previously linked to AD, but their measurement in plasma was hitherto not established. Our findings, therefore, pave the way for independent validation of these plasma markers to enable their widespread use for diagnostic, prognostic, and monitoring.

## Abbreviations

Aβ: Amyloid-beta
AD: Alzheimer’s disease
AUC: Area under Curve
BBB: Blood brain barrier
CDR: Clinical dementia rating
CNS: Central nervous system
CSF: Cerebrospinal fluid
CV: Coefficient of variation
FDR: False discovery rate
IC: Internal control
IQR: Interquartile range<colcnt=2>
LOD: Limit of detection
MCI: Mild cognitive impairment
MMSE: Mini-Mental State Examination
MRI: Magnetic resonance imaging
MYHAT: Monongahela Youghiogheny Healthy Aging Team
MYHAT: NI Monongahela Youghiogheny Healthy Aging Team-Neuroimaging
NPQ: NULISA protein quantification
NULISA: NUcleic acid-linked Immuno-Sandwich Assay
NULISAseq: NULISA with next-generation sequencing readout
PiB: Pittsburgh compound-B
PLA: Proximity ligation assay
p-tau: Phosphorylated tau
PET: Positron emission tomography
QC: Quality control
ROC: Receiver operating characteristic
ROIs: Regions of interest
SD: Standard deviation
Simoa: Single-molecule array
SUVR: Standardized uptake value ratio

## Acknowledgments

We thank all members of the Karikari Laboratory and Dr. Rebecca Deek for statistical advice. We are indebted to the participants, family members, and staff of the MYHAT-NI study.

## Author contributions

TKK, MIK, ADC, BES, and MG contributed to the study’s conception and design. BES and MG ran MYHAT and MYHAT-NI cohorts. ADC, VLV, TAP, PCLF, BB, GP, OIL and WEK contributed to neuroimaging data collection and analysis. TKL, AS, PCF, BB, and PG performed biochemical assays. MIK performed genotyping experiments. XZ and YC performed data analysis and produced the figures. XZ and TKK were major contributors in writing the manuscript. All authors contributed to and approved the final version of the manuscript.

## Funding

TKK was supported by the NIH (R01 AG083874, U24 AG082930, P30 AG066468, RF1 AG052525-01A1, R01 AG053952-05, R37 AG023651-17, RF1 AG025516-12A1, R01 AG073267-02, R01 AG075336-01, R01 AG072641-02, P01 AG025204-16) and the Alzheimer’s Association (#AARF-21-850325). MDI was supported by NIH/NIA grants P01AG14449 and P01AG025204. The MYHAT study was supported by R37 AG023651-17 and MYHAT-NI by R01 AG052521.

## Data availability

De-identified, cohort-level data will be shared at the request of verified investigators to replicate procedures and results reported in this article. Data transfer agreements in accordance with US legislation and the decisions of the University of Pittsburgh’s Institutional Review Board, which covers the jurisdiction of the MYHAT-NI study, may need to be established.

## Declarations

### Ethics approval and consent to participate

All plasma samples were obtained with full written informed consent and approved by the University of Pittsburgh Institutional Review Board (STUDY19020264).

### Consent for publication

Not applicable.

### Competing interest

The authors declare that they have no competing interests

## Supplementary Information

Additional file 1: Supplementary Figure S1 to S5

## Tables

**Table 1:** Participant characteristics in the MYHAT-NI cohort

**Table 2:** Diagnostic accuracy (ROC analysis) of NULISAseq and Simoa biomarkers for Aβ PET positivity.

## Notes

### Competing Interest Statement

The authors have declared no competing interest.

### Author Declarations

The MYHAT-NI study was approved by the University of Pittsburgh Institutional Review Board (STUDY19020264).

