## Supplementary Figures S1-S5 for "Multi-analyte proteomic analysis identifies blood-based neuroinflammation, cerebrovascular and synaptic biomarkers in preclinical Alzheimer’s disease"

**Additional file 1**

**Figure S1** NULISAseq targets with significant association with common risk factors (age, sex, and APOE ε4 carrier status).

**Figure S2** Association between longitudinal changes in Aβ PET SUVR and NULISAseq targets.

**Figure S3** Receiver operating characteristics (ROC) curves for p-tau217 and p-tau231 in detecting tau PET positivity.

**Figure S4** Longitudinal association between NULISAseq targets and tau pathology (T).

**Figure S5** Longitudinal association between NULISAseq targets and neurodegeneration (N).

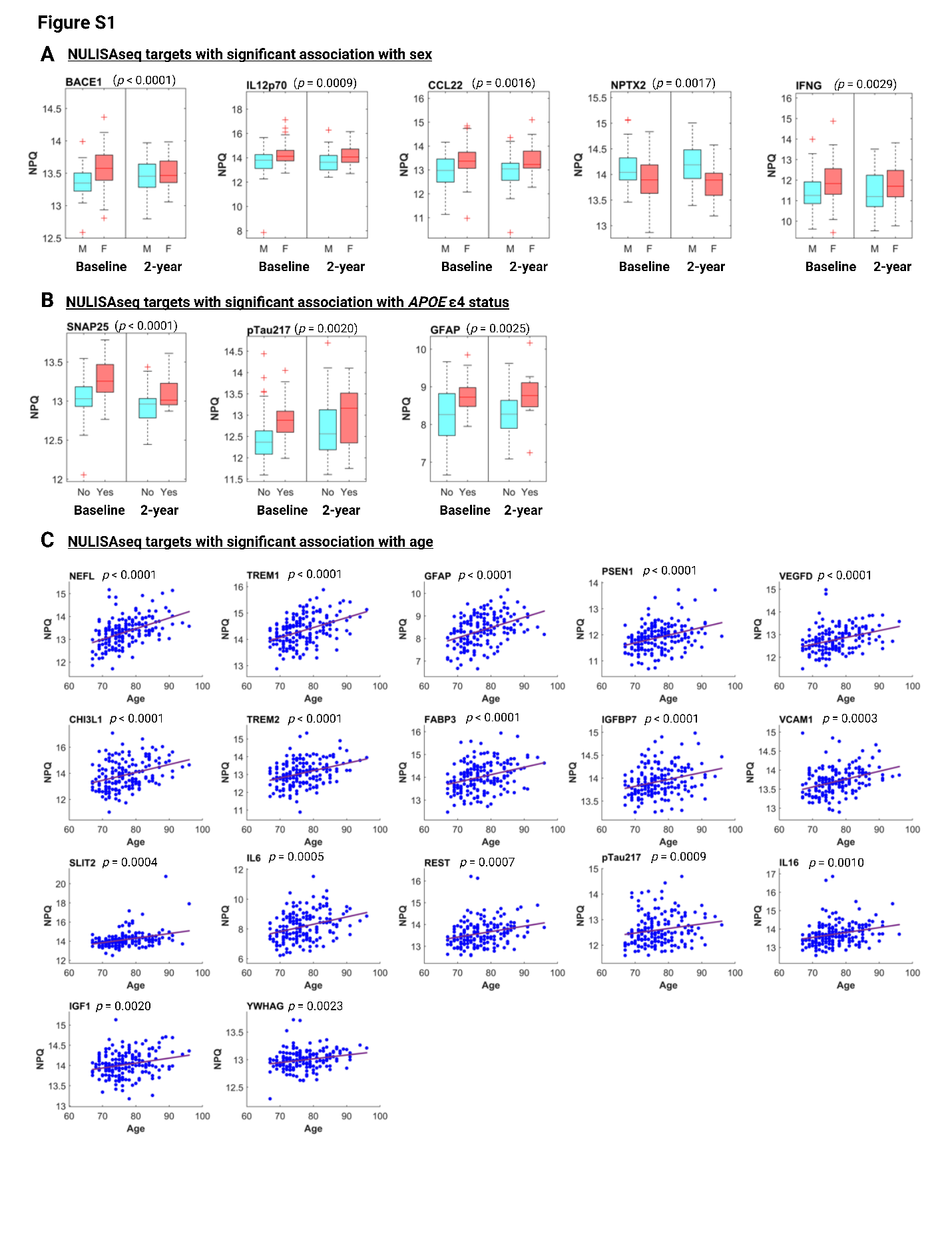

**Figure S1** NULISAseq targets with significant association with common risk factors. **A** Boxplots of NULISAseq targets with significant association with biological sex, separated by sex and visit. M refers to males, and F refers to females. **B** Boxplots of NULISAseq targets with significant association with *APOE* ε4 carrier status, separated by *APOE* ε4 carrier status and visit. The participants with at least one *APOE* ε4 allele were grouped as “Yes” and those with no *APOE* ε4 as “No.” **B** Scatterplots illustrating the correlation between age and NULISAseq levels. P-values on top of all boxplots were for the whole data combining both visits and were determined using linear mixed models (random intercepts) with NPQs as the dependent variable and the corresponding covariates as the independent variables. Significance determination was based on p-value < 0.005, corresponding to 10%, 10%, and 3% FDR for sex, *APOE* ε4 carrier status, and age, respectively. Purple lines indicated the least square regression lines.

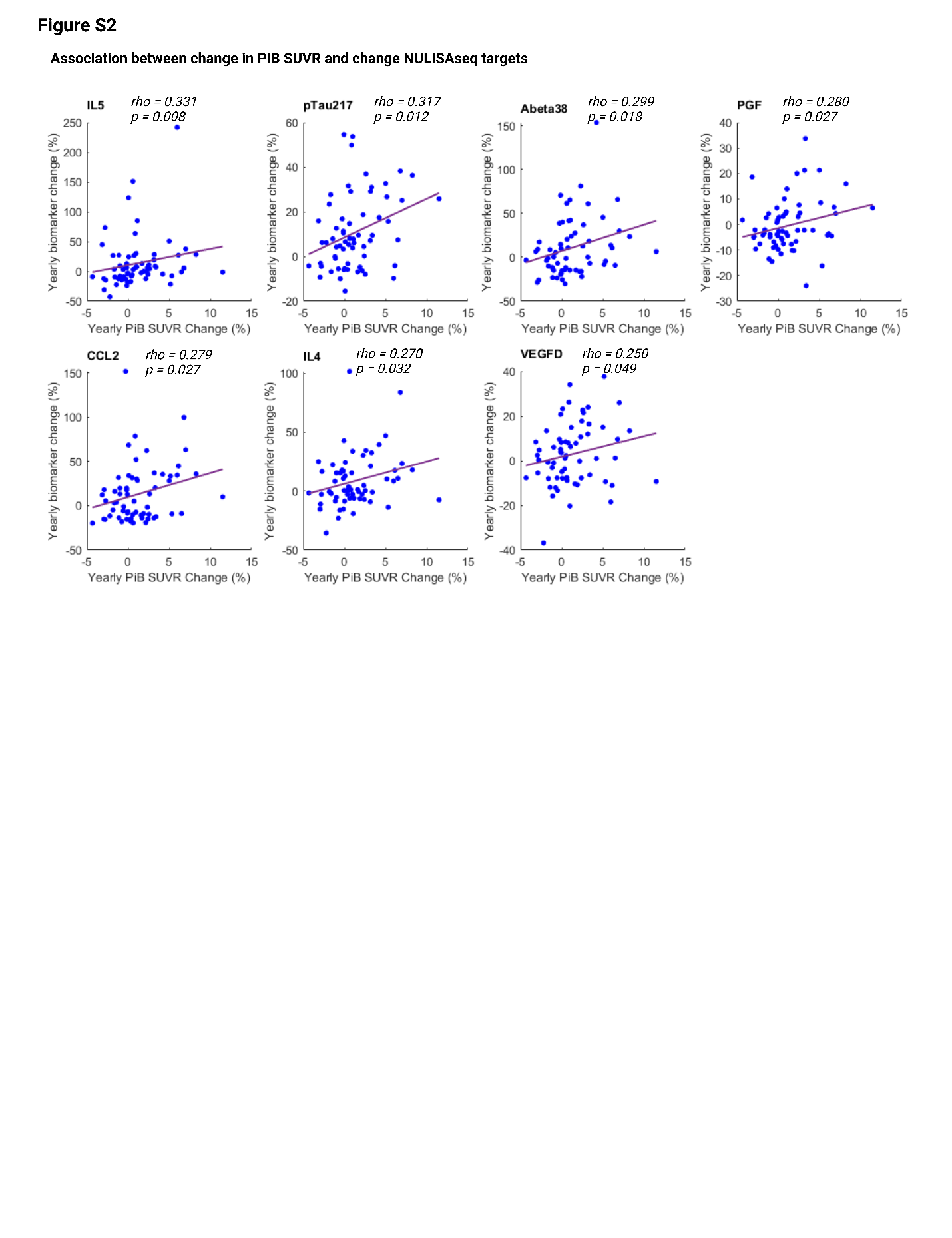

**Figure S2** NULISAseq targets with significant associations between longitudinal changes in Aβ PET SUVR and biomarker levels. Shown are scatterplots between yearly Aβ PET SUVR change (x-axis) and yearly biomarker level change (y-axis). The strength of the correlation was assessed using Spearman’s ranks. Purple lines indicated the least square regression lines. The plasma biomarkers are arranged in decreasing order of the strength of association.

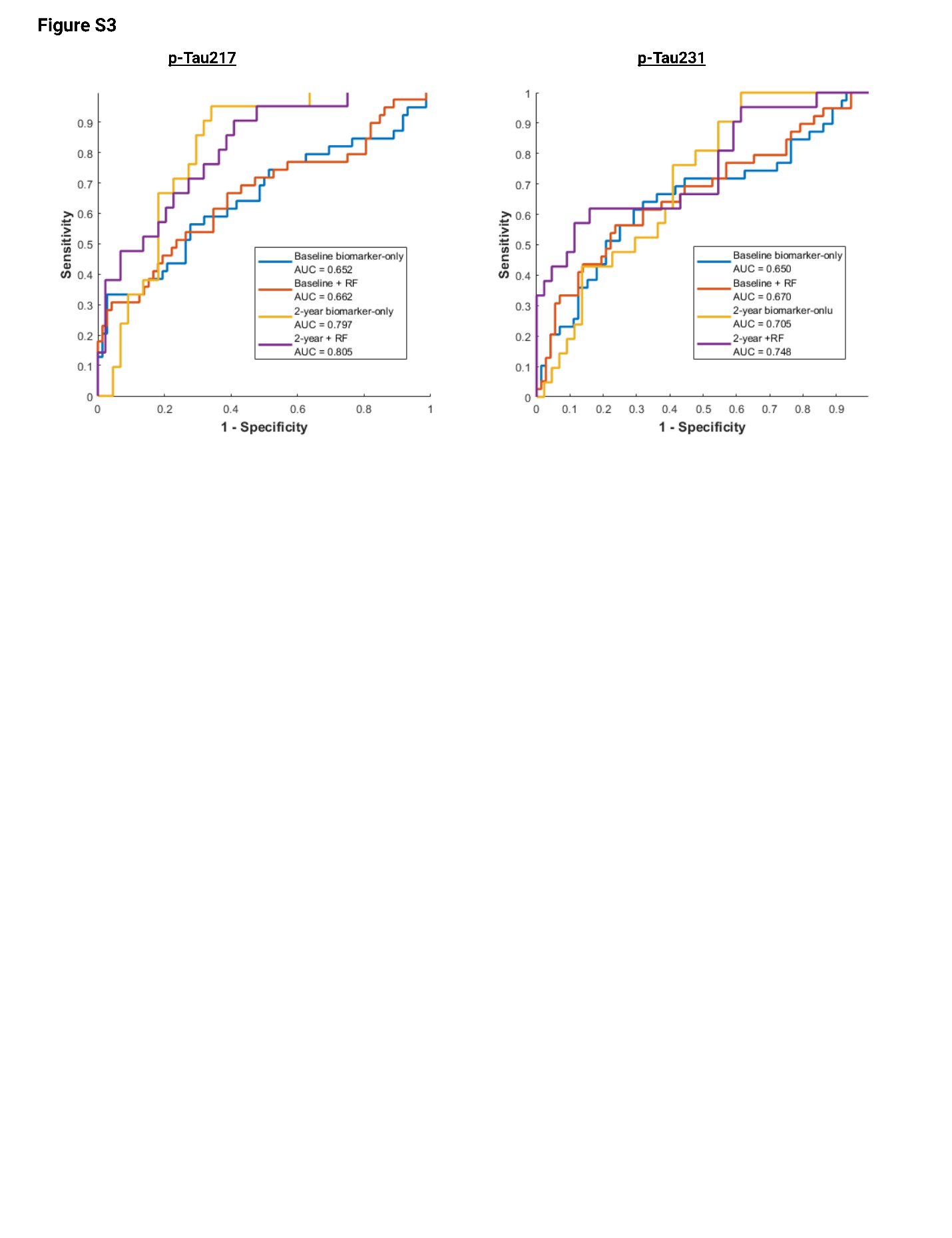

**Figure S3** Receiver operating characteristics (ROC) curves for p-tau217 and p-tau231 in detecting tau PET positivity. RF refers to a model incorporating age, sex, and *APOE* ε4 carrier status. ROC curves and the corresponding area under the curve (AUC) were determined using the MATLAB *perfcurve* function.

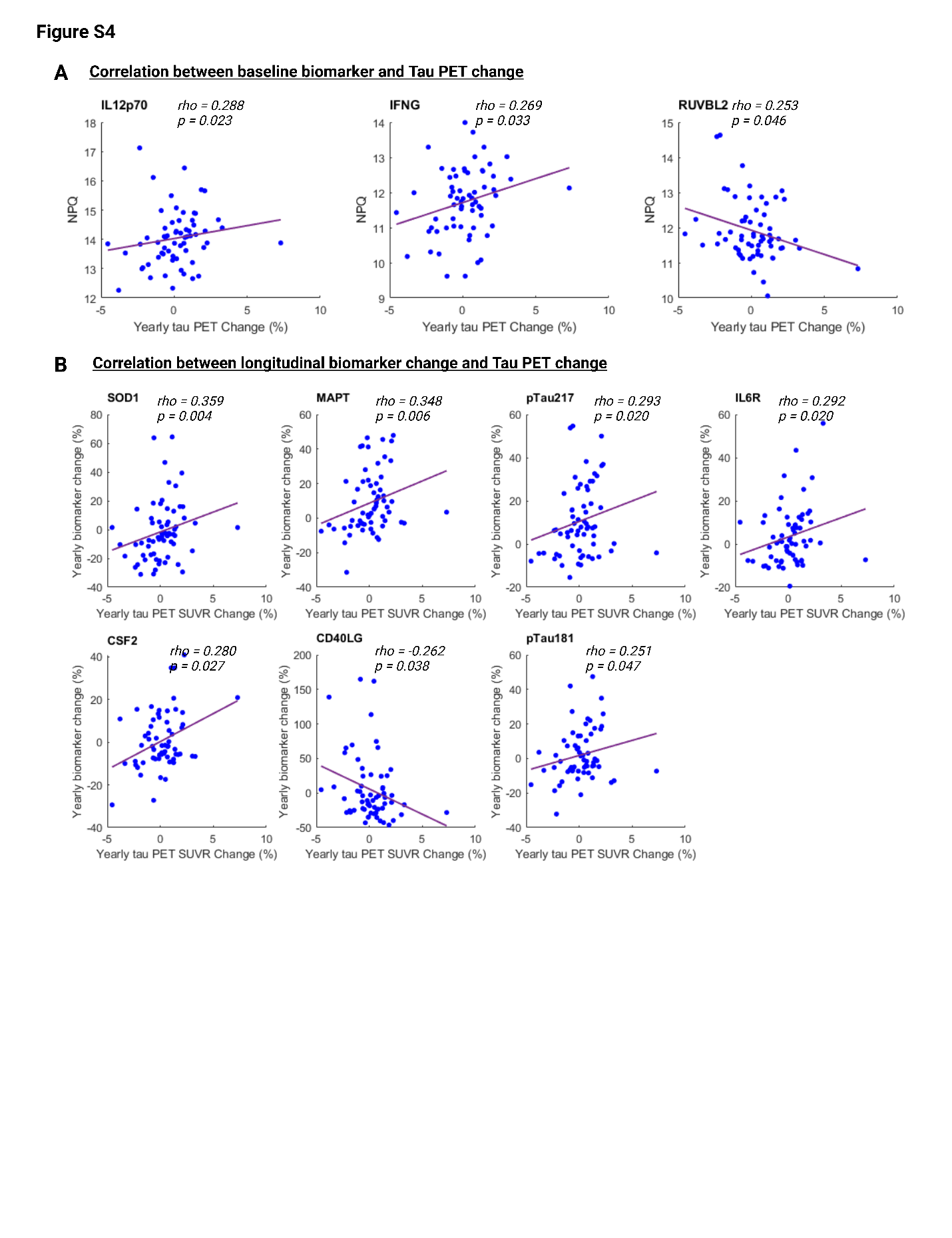

**Figure S4** Longitudinal association between NULISAseq targets and tau pathology (T). **A** Scatterplots depicting the correlation between yearly longitudinal tau PET SUVR change (x-axis) and baseline biomarker levels (y-axis). **B** Scatterplots illustrating the correlation between yearly longitudinal tau PET SUVR change (x-axis) and yearly biomarker level change (y-axis). All correlations were based on Spearman's rank correlation. Purple lines indicated the least square regression lines.

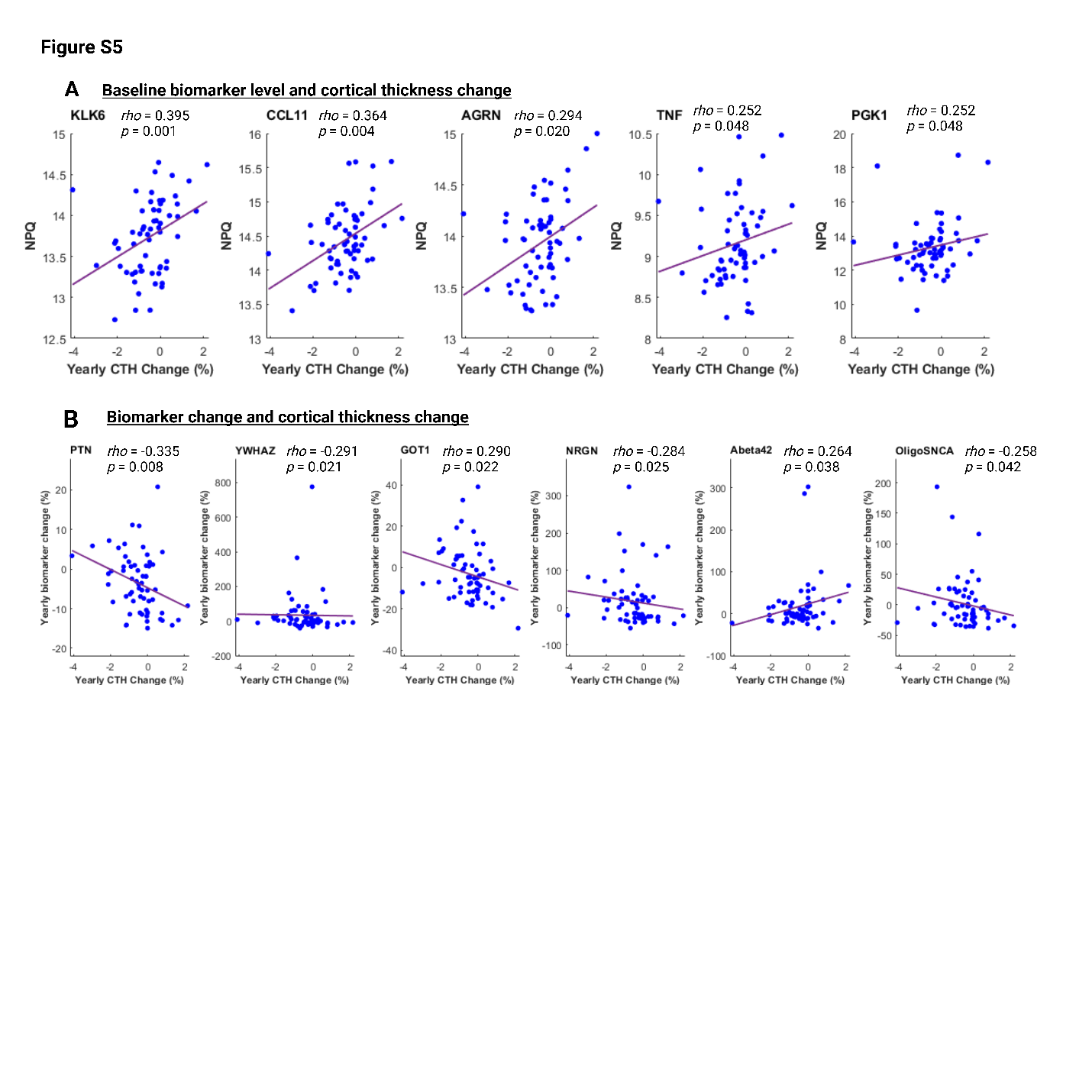

**Figure S5** Longitudinal association between NULISAseq targets and neurodegeneration (N). **A** Scatterplots depicting the correlation between yearly longitudinal MRI-determined cortical thickness change (CTH; x-axis) and baseline biomarker levels (y-axis). **B** Scatterplots illustrating the correlation between yearly CTH change (x-axis) and yearly biomarker level change (y-axis). All correlations were based on Spearman's rank correlation. Purple lines indicated the least square regression lines.
